# Ad26.COV2.S breakthrough infections induce high titers of neutralizing antibodies against Omicron and other SARS-CoV-2 variants of concern

**DOI:** 10.1101/2021.11.08.21266049

**Authors:** Dale Kitchin, Simone I. Richardson, Mieke A. van der Mescht, Thopisang Motlou, Nonkululeko Mzindle, Thandeka Moyo-Gwete, Zanele Makhado, Frances Ayres, Nelia P. Manamela, Holly Spencer, Bronwen Lambson, Brent Oosthuysen, Haajira Kaldine, Marizane du Pisanie, Mathilda Mennen, Sango Skelem, Noleen Williams, Ntobeko A.B. Ntusi, Wendy A. Burgers, Glenda G. Gray, Linda-Gail Bekker, Michael T. Boswell, Theresa M. Rossouw, Veronica Ueckermann, Penny L. Moore

## Abstract

The Janssen (Johnson & Johnson) Ad26.COV2.S non-replicating viral vector vaccine has been widely deployed for COVID-19 vaccination programs in resource-limited settings. Here we confirm that neutralizing and binding responses to Ad26.COV2.S vaccination are stable for 6 months post-vaccination, when tested against multiple SARS-CoV-2 variants. Secondly, using longitudinal samples from individuals who experienced clinically mild breakthrough infections 4 to 5 months after vaccination, we show dramatically boosted binding antibodies, Fc effector function and neutralization. These high titer responses are of similar magnitude to humoral immune responses measured in severely ill, hospitalized donors, and are cross-reactive against diverse SARS-CoV-2 variants, including the extremely neutralization resistant Omicron (B.1.1.529) variant that currently dominates global infections, as well as SARS-CoV-1. These data have implications for population immunity in areas where the Ad26.COV2.S vaccine has been widely deployed, but where ongoing infections continue to occur at high levels.

## Main Text

A phase 3 clinical trial of Ad26.COV2.S in eight countries demonstrated 85% protection against severe disease^1^, including in South Africa, where the trial coincided with the emergence of the Beta (B.1.351) variant, which was shown to have increased resistance to neutralizing antibodies^2,3^. As a result, Ad26.COV2.S was made available to South African health care workers (HCWs) in early 2021 through the Sisonke open-label, phase 3b clinical trial. Globally, this vaccine has also been used widely in several countries, including USA, and EU member states, with 5.38, 15.68 and 16.16 million doses administered in these regions, respectively, by the beginning of November 2021.

Subsequently, South Africa has experienced a third and fourth wave of infection, driven by the Delta (B.1.617.2) and Omicron (B.1.1.529) variants, respectively, with reports of Ad26.COV2.S breakthrough infections (BTIs) occurring. Infections following mRNA vaccination result in boosted neutralizing antibody titers^4–6^, but less is known about the immunological consequences of BTI after Ad26.COV2.S vaccination. With the emergence in late November 2021^7^ of the Omicron variant which now dominates global infections and has more spike mutations in key neutralizing epitopes than any variant to date, a key question is whether Ad26.COV2.S vaccinated individuals who experienced breakthrough infections in the previous Delta-driven wave would have substantial neutralizing responses against this variant.

Here, we evaluated the durability and breadth of vaccine-elicited humoral responses in nineteen HCWs vaccinated with Ad26.COV.2S in February-March 2021 (Fig. 1a). Secondly, we characterised the humoral response to BTI in a subset of six individuals with SARS-CoV-2 PCR-confirmed infections 4-5 months (median number of months: 4.4; interquartile range: 4.1-4.8) following vaccination. Five of these participants were followed longitudinally 2-6 months post-vaccination, while for the sixth BTI participant only a single sample from 1 month post-infection was available (Table S1). These BTIs occurred between June and August 2021, during the third wave of SARS-CoV-2 infections in South Africa. This wave was driven by the more transmissible Delta variant, which accounted for between 40% and 95% of genomes sequenced in South Africa over this period^8^. BTIs were thus most likely caused by the Delta variant, though sequencing data for these participants were not available. Participants, of whom 16/19 were female, had a median age of 34 (interquartile range: 30-40 years) and all presented with mild disease (Table S1). All nineteen participants were SARS-CoV-2 naive prior to vaccination, as confirmed by nucleocapsid ELISA (Fig. 1b).

**Fig. 1:**
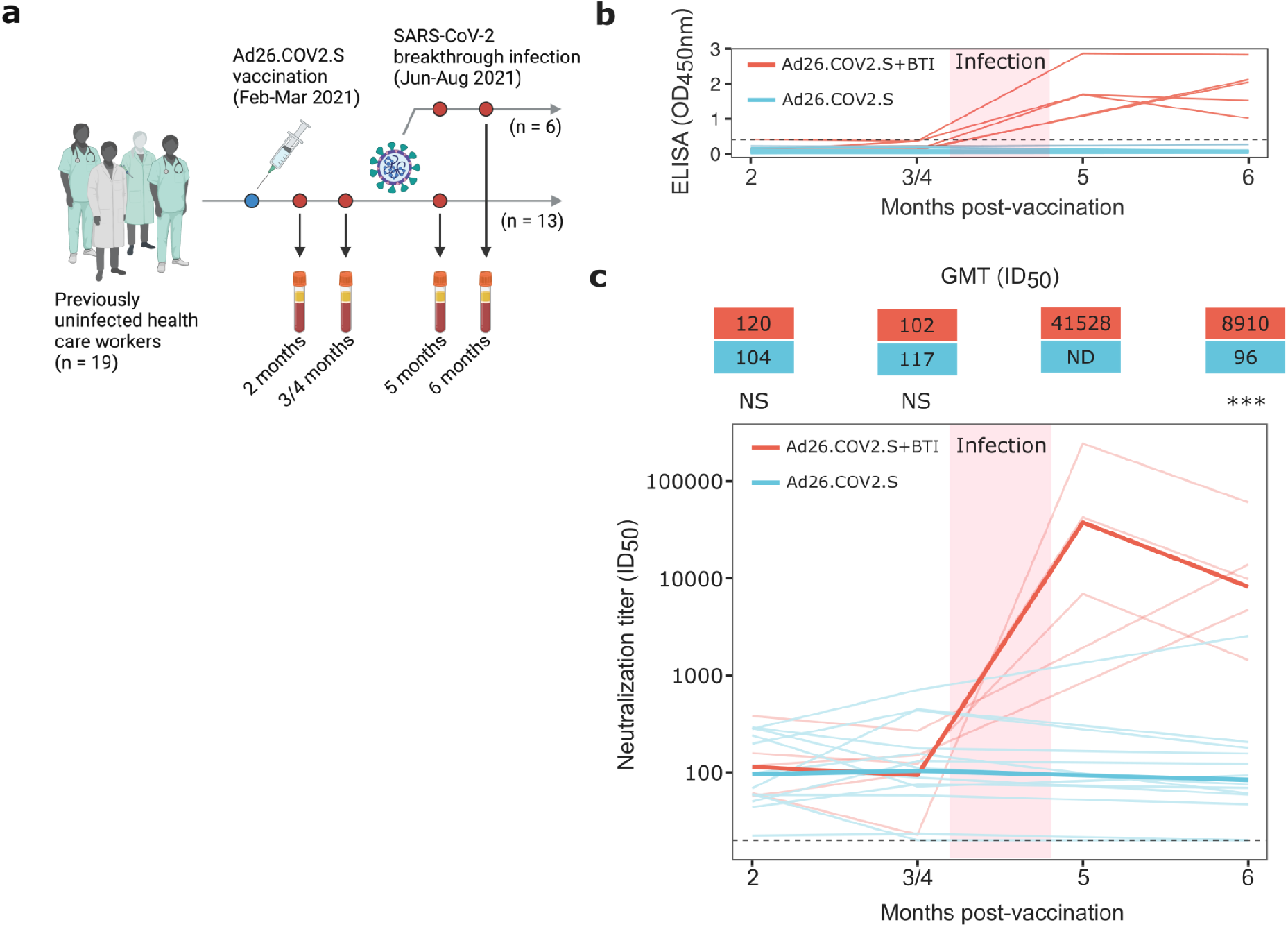
Ad26.COV2.S breakthrough infection (BTI) results in boosted plasma neutralization titers against the ancestral variant (D614G). **(a)** Nineteen South African HCWs, vaccinated between February and March 2021 with a single dose of Ad26.COV2.S, were recruited into a vaccine durability study, with longitudinal blood draws occurring at 2-, 3- (or 4-), 5- and 6-months post-vaccination. Six of the HCWs had BTIs (PCR-confirmed) between June and August 2021. **(b)** Nucleocapsid ELISA binding (OD_450nm_), from 2-6-months post-vaccination, is shown for each BTI and non-BTI participant by red and blue lines, respectively. The threshold of positivity is indicated by the dashed horizontal line. **(c)** Neutralization titers (ID_50_) against the D614G variant, from 2-6-months post-vaccination, are shown for each BTI and non-BTI participant by red and blue lines, respectively. Lines in bold indicate the GMTs for the BTI and non-BTI groups, with the values indicated above the figure in the blue and red boxes. The horizontal dashed line indicates the lower limit of quantitation. Statistical analyses were performed using the Mann-Whitney test between groups, with *** denoting p<0.001, NS for non-significant and ND for no data.

We first assessed the durability of vaccine-elicited antibody responses in individuals who were confirmed to have remained uninfected up to 6-months post vaccination, by nucleocapsid ELISA (Fig. 1b). Spike binding responses against the original D614G variant were measured at 2-, 4- and 6-months post-vaccination. No significant reduction in binding was observed over this period (Fig. S1a), as has been previously reported^9–11^. We also used a spike-pseudotyped lentivirus neutralization assay, to measure longitudinal neutralization titers against the ancestral D614G variant (which differs from the vaccine spike protein by a single D614G mutation), six SARS-CoV-2 variants with increased transmissibility and/or immune escape mutations, namely Beta, Delta, Gamma (P.1), C.1.2, A.VOI.V2, and Omicron as well as SARS-CoV-1. For the ancestral D614G variant, geometric mean titers (GMT) were stable up to 6 months post-vaccination (GMTs of 104, 117 and 96 at 2-, 4- and 6-months post-vaccination), consistent with previous studies^10,11^ (Fig. 1c, S2a). Where detectable, titers against the six variants were similarly stable over 6 months, showing no significant differences over time (Fig. S2a), as observed in two previous studies^10,12^. However, for all variants tested, titers were 1.9-4.2-fold lower at 2-months post-vaccination compared to the D614G variant, as reported elsewhere^10,12–15^ (Fig. S2a). For the Beta and Delta variants, in particular, half of non-BTI vaccinees showed no detectable neutralization at 6-months post-vaccination (Fig. S2c). As expected, titers against SARS-CoV-1 were low with GMTs of 28 and 21 at 2- and 4-months respectively, and undetectable at 6-months post-vaccination (Fig. S2a).

We next assessed the breadth and magnitude of humoral immune responses following BTI. In all participants, BTI occurred between 3- and 5-months post-vaccination. Prior to BTI, the nucleocapsid binding responses in both the BTI and non-BTI participants were negative, and only detected following BTI. (Fig 1b). There were also no significant differences in D614G spike binding responses between the BTI and non-BTI participants prior to 3- to 4-months post-vaccination (Fig. S1a). However, following infection, there was a 3.3-fold increase in spike responses, which peaked at approximately 2-weeks post infection (5-months post-vaccination) and remained constant until 1-month post-infection (6-months post-vaccination) (Fig. S1a).

We also assessed the impact of BTI on Fc effector functions, which have been implicated in protection from severe COVID-19 disease, and which generally retain activity against variants of concern (VOCs)^16,17^. We examined whether levels of antibody-dependent cellular cytotoxicity (ADCC), measured by ability to cross-link FcγRIIIa were boosted following BTI. Similar to binding, ADCC against D614G remained stable up to 3- to 4-months post-vaccination, with a rapid 3.1-fold increase in activity after BTI (Fig. S1b). These responses peaked (geomean RLU: 712) at approximately 2-weeks post-infection (5-months post-vaccination), but declined slightly (geomean RLU: 558) by 1-month post-infection (6-months post-vaccination) (Fig. S1b). Both before and after infection, ADCC was cross-reactive against D614G, Beta and Delta variants, showing only slight decreases against VOCs relative to D614G across all time points (geomean RLUs of 712, 626 and 702 against D614G, Beta and Delta, respectively, at 2-weeks post-infection). This illustrates the resilience of Fc effector function against VOCs in Ad26.CoV2.S BTI participants.

Neutralization titers against D614G closely mirrored the spike binding and ADCC response, with no significant differences in titers between the BTI and non-BTI participants prior to 3- to 4-months post-vaccination, but with a dramatic increase in titers for all participants (407-fold increase from 102 to 41528 GMT) following infection (Fig. 1c). This increase in neutralization titers is similar in magnitude to what was previously reported for a single individual with Ad26.CoV2.S BTI^10^. Neutralization titers peaked at approximately 2 weeks post-infection (5-months post-vaccination) and declined by approximately 4.7-fold one month thereafter (Fig. 1c). Neutralization titers after BTI were also significantly higher against six SARS-CoV-2 variants relative to non-BTI participants (40-154-fold difference in GMT), and SARS-CoV-1 (9-fold difference in GMT) (Fig. 2, S2b). This includes the highly neutralization resistant Omicron variant, against which titers ranged from 161 to 1858 (GMT of 843) for the BTI participants (Fig. 2). Thus, in contrast to vaccine-elicited responses, BTI after a single dose of Ad26.COV2.S resulted in complete coverage of SARS-CoV-2 variants other than Omicron, with GMTs >1:3,000 (Fig. 2, S2a, S2b). For Omicron, while all BTI participants had detectable titers, these were dramatically lower than for all other variants, and more comparable in titer to those observed against SARS-CoV-1(Fig. 2).

**Fig. 2:**
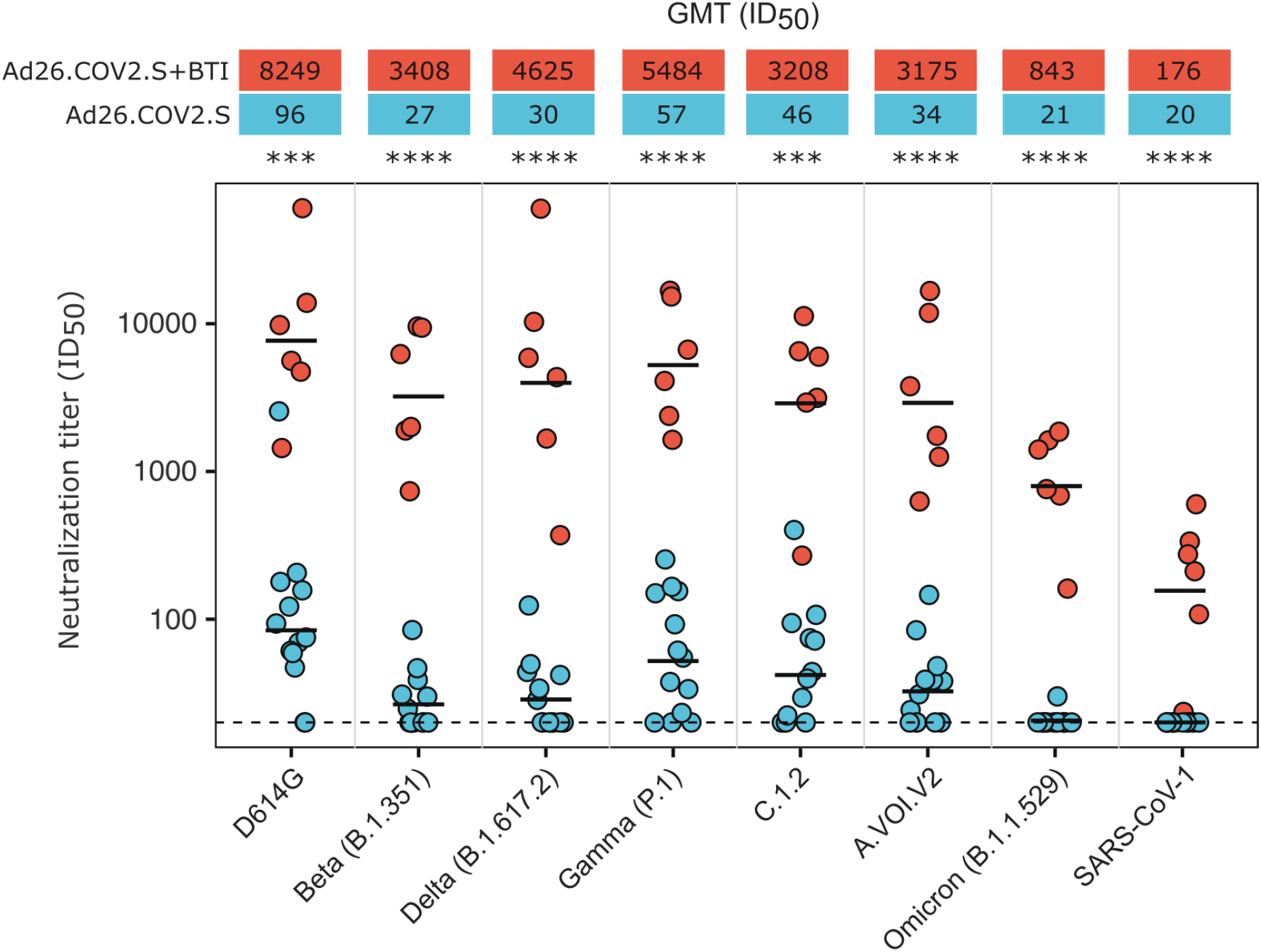
Breakthrough infection (BTI) results in increased plasma neutralization titers against all SARS-CoV-2 variants, and SARS-CoV-1, 6-months post-vaccination. The neutralization titers against the ancestral (D614G), Beta (B.1.351), Delta (B.1.617.2), Gamma (P.1), C.1.2, A.VOI.V2 and Omicron (B.1.1.529) SARS-CoV-2 variants, and SARS-CoV-1, for six BTI participants relative to thirteen non-BTI participants at the 6-months post-vaccination visit (approximately 1-month post-BTI). Each dot represents the neutralization titer of a single participant, with the BTI participants and non-BTI participants shown in red and blue, respectively. The GMT for each group against each variant is shown by a black horizontal bar in the plot, with the values given in the red and blue boxes above the plot. Neutralization titers in the BTI group were significantly higher than those of the non-BTI group (40 to 154-fold higher GMT against the SARS-CoV-2 variants and 9-fold higher against SARS-CoV-1). Statistical analyses were performed using the Mann-Whitney test between groups, with *** denoting p<0.001 and **** denoting p<0.0001.

Overall our data confirms durable vaccine-elicited humoral immune responses 6 months after a single dose of Ad26.COV2.S, consistent with other studies10,12–15. Moreover, despite relatively modest titers after vaccination, we observe significantly boosted binding antibodies, ADCC and neutralization activity following BTI. This boost resulted in neutralization titers in BTI participants at 1-month post-infection (GMT 8,249) that were higher than those elicited by a 2 dose Pfizer-BioNTech (BNT162b2) vaccine regimen (GMT: 1,128) 2-months post vaccination, or those observed in acutely infected hospitalised individuals with moderate (GMT: 993) or severe disease (GMT: 3,747) (Fig. S3). Though we note that these comparisons differ by number of antigen exposures and timing, amongst other variables, this illustrates the extremely high level of boosting observed in BTIs. Similar to BTI individuals, we have previously confirmed that ADCC, binding and neutralization are also significantly boosted following vaccination in individuals who were previously infected18, but not to the same levels we report here for BTI (1,372 vs 8,249 for neutralization, respectively) (Fig. S3). Whether this is a result of increased affinity maturation broadening the response against variants, or rather the boosting of vaccine elicited cross-reactive memory B cells to higher titres, remains to be determined and will be the focus of future work.

These data add to previous reports of BTI following mRNA vaccination which results in >30-fold increased neutralization potency, suggesting broad relevance across multiple vaccine modalities^5,19^. Taken together, these findings suggest a strongly synergistic effect of vaccination and infection which will contribute to higher levels of protective community immunity in areas with high burden of infection, and may be contributing to the low hospitalisation rates seen during the current Omicron-driven fourth wave in South Africa^20^. As homologous and heterologous boosters are deployed this effect may be further enhanced^21^. Overall, this study provides insight into the magnitude and quality of humoral immune responses elicited by breakthrough infections after an adenovirus-based vaccine, with implications for public health interventions in regions that have experienced high levels of SARS-CoV-2 transmission.

## Limitations of study

This study is limited by the relatively small number of BTI that were characterised. We also note that the median age of the BTI participants was higher than that of the non-BTI participants (BTI participants median age: 39 years, interquartile range: 32-59 years; non-BTI participants median age: 33 years, interquartile range: 29-36 years) which may impact comparisons between the two groups^11,22^. We do not have sequencing data to determine the variant responsible for BTI, and it is possible that the infecting viral sequence may impact the quality of the response, as we have previously shown^18,23^. We note that the pseudotyped neutralization assay used here can only detect neutralizing antibodies against the spike, the primary target for these antibodies, unlike the live neutralization assay which can also detect those directed at the nucleocapsid, and we may therefore be underestimating overall immunity. We also focused only on humoral responses, and T cell responses following BTI will need to be defined in future studies.

## Data Availability

All data produced in the present study are available upon reasonable request to the authors.

## Acknowledgements

We thank the study participants at Groote Schuur Hospital, Steve Biko Academic Hospital and the National Institute for Communicable Diseases, who contributed samples that enabled this work. We also thank Elloise du Toit for assistance with clinical data and Nigel Garrett, Ameena Goga and the Sisonke vaccination team. Tandile Hermanus assisted with the analysis of neutralization data. Roanne Keeton, Amkele Ngomti, Richard Baguma and Ntombi Benede assisted with sample processing and storage. The parental soluble spike was provided by Jason McLellan (University of Texas). The parental pseudovirus plasmids were kindly provided by Elise Landais and Devin Sok (IAVI).

## Author contributions

D.K., S.I.R. and P.L.M. conceived the study, designed experiments, analyzed data and wrote the paper. Z.M., F.A., B.O. and B.E.L. made molecular constructs and expressed antibodies. Z.M. and T.M.G. expressed and purified recombinant antigens. Z.M. and F.A. performed spike and nucleocapsid ELISAs. T.M., N.M. and H.K. made pseudoviruses and performed neutralization experiments. S.I.R., N.P.M and H.S. performed ADCC assays. L.G.B and G.G.G conceptualized and led the Sisonke Ad26.COV2.S trial. S.I.R. established the NICD HCW cohort of vaccinees. M.A.v.d.M, M.d.P, M.T.B, T.M.R., V.U., established the Steve Biko HCW cohort and provided samples. M.M., S.S, N.W., N.A.B.N. and W.A.B. established the Groote Schuur Hospital cohort and provided samples. All authors critically reviewed and approved the final manuscript.

## Funding

This work was supported by the South African Medical Research Council (grants 96825 and 96838). P.L.M. is supported by the South African Research Chairs Initiative of the Department of Science and Innovation and the National Research Foundation of South Africa (grant no. 98341). W.A.B. is supported by the EDCTP2 programme of the European Union’s Horizon 2020 programme (TMA2016SF-1535-CaTCH-22), the Wellcome Centre for Infectious Diseases Research in Africa (CIDRI-Africa) which is supported by core funding from the Wellcome Trust (203135/Z/16/Z) and the Poliomyelitis Research Foundation (PRF 21/65). N.A.B.N acknowledges funding from the SA-MRC, MRC UK, NRF and the Lily and Ernst Hausmann Trust. S.I.R. is a L’Oreal/UNESCO Women in Science South Africa Young Talents awardee. Related research by the authors is conducted as part of the DST-NRF Centre of Excellence in HIV Prevention, which is supported by the Department of Science and Technology and the National Research Foundation.

## Materials and methods

### Human samples

HCWs vaccinated with one dose of Ad26.CoV2.S (5×10^10^ viral particles) as part of the Sisonke implementation trial were followed longitudinally and plasma sampled at 2-, 3- (in some cases 4-) and 6-months post-vaccination. An additional plasma sample was collected from BTI participants at 5-months post-vaccination, which was approximately 2-weeks post-infection. Non-BTI participants were recruited from HCWs at the National Institute for Communicable Diseases (NICD) (Johannesburg), while BTI participants were recruited from HCWs at the NICD, Steve Biko Academic Hospital (Tshwane, South Africa) and Groote Schuur Hospital (Cape Town, South Africa). Ad26.CoV2.S vaccinees with prior SARS-CoV-2 infection were recruited from a longitudinal study of healthcare workers enrolled from Groote Schuur Hospital, with plasma samples collected 2-months post-vaccination. Plasma was also collected from thirteen participants that had received two doses of the Pfizer BioNTech vaccine (BNT162b2) 2-months after they had received their last dose (Johannesburg, South Africa). Convalescent participants were recruited as part of a hospitalised cohort at the Steve Biko Academic Hospital between May and August 2020, with plasma samples collected 10-days after the initial positive PCR test. Ethics approval was obtained from the Human Research Ethics Committees of the University of the Witwatersrand (ethics reference number: M210465), University of Pretoria (ethics reference number: 247/2020) and University of Cape Town (ethics reference numbers: 190/2020 and 209/2020). Written informed consent was obtained from all participants.

### SARS-CoV-2 antigens

For ELISA, SARS-CoV-2 original variant (D614G) full spike proteins were expressed in Human Embryonic Kidney (HEK) 293F suspension cells by transfecting the cells with the respective expression plasmid. After incubating for six days at 37°C, 70% humidity and 10% CO_2_, proteins were first purified using a nickel resin, followed by size-exclusion chromatography. Relevant fractions were collected and frozen at −80°C until use. A commercial, recombinant nucleocapsid protein (BioTech Africa, Cat. no.: BA25-C) was used as the antigen in the nucleocapsid ELISAs.

### SARS-CoV-2 Spike and nucleocapsid enzyme-linked immunosorbent assay (ELISA)

Spike or nucleocapsid protein (2 μg/mL) was used to coat 96-well, high-binding plates and incubated overnight at 4°C. The plates were incubated in a blocking buffer consisting of 5% skimmed milk powder, 0.05% Tween 20, 1x PBS. Plasma samples were diluted to 1:100 starting dilution in a blocking buffer and added to the plates. IgG secondary antibody was diluted to 1:3000 or 1:1000 respectively in blocking buffer and added to the plates followed by TMB substrate (Thermo Fisher Scientific). Upon stopping the reaction with 1M H_2_SO_4_, absorbance was measured at a 450 nm wavelength. In all instances, mAbs CR3022 and BD23 were used as positive controls and palivizumab was used as a negative control.

### Spike plasmid and lentiviral pseudovirus production

The SARS-CoV-2 Wuhan-1 spike gene sequence, cloned into pcDNA3.1, was mutated using the QuikChange Lightning Site-Directed Mutagenesis kit (Agilent Technologies) and NEBuilder HiFi DNA Assembly Master Mix (New England Biosciences) to include D614G (ancestral) or lineage defining mutations for Beta (L18F, D80A, D215G, Δ242-244, K417N, E484K, N501Y, D614G and A701V), Delta (T19R, Δ156-157, R158G, L452R, T478K, D614G, P681R, D950N), Gamma (L18F, T20N, P26S, D138Y, R190S, K417T, E484K, N501Y, D614G, H655Y, T1027I, V1176F), C1.2. (P9L, C136F, Δ144, R190S, D215G, Δ242-243, Y449H, E484K, N501Y, D614G, H655Y, N679K, T716I, T859N), A.VOI.V2 (D80Y, Δ144, I210N, Δ211, D215G, R246M, Δ247-249, W258L, R346K, T478R, E484K, H655Y, P681H, Q957H) and Omicron (A67V, Δ69-70, T95I, G142D, Δ143-145, Δ211, L212I, 214EPE, G339D, S371L, S373P, S375F, K417N, N440K, G446S, S477N, T478K, E484A, Q493K, G496S, Q498R, N501Y, Y505H, T547K, D614G, H655Y, N679K, P681H, N764K, D796Y, N856K, Q954H, N969K, L981F). SARS-CoV-1 spike was also cloned into pcDNA3.1. Pseudotyped lentiviruses were prepared as previously described3. Briefly, 293T/17 cells (HEK293T cell line) were co-transfected with a SARS-CoV-2 spike plasmid in conjunction with a firefly luciferase encoding lentivirus backbone plasmid (pNL4-3.Luc.R-E-) with PEI MAX (Polysciences). Culture supernatants were clarified of cells by a 0.45-μM filter and stored at −70°C.

### Pseudovirus neutralization assay

For the neutralization assay, plasma samples were heat-inactivated and clarified by centrifugation. Heat-inactivated plasma samples from vaccine recipients were incubated with the SARS-CoV-1/2 pseudotyped virus for 1 hour at 37°C, 5% CO_2_. Subsequently, 1×10^4^ HEK293T cells engineered to over-express ACE-2 (293T/ACE2.MF), kindly provided by M. Farzan (The Scripps Research Institute) were added and incubated at 37°C, 5% CO_2_ for 72 hours upon which the luminescence of luciferase was measured. Titers were calculated as the reciprocal plasma dilution (ID_50_) causing 50% reduction of relative light units (RLU). Monoclonal antibodies CB6 and CA1 were used as positive controls.

### Antibody-dependent cellular cytotoxicity (ADCC) assay

The ability of plasma antibodies to cross-link spike expressing cells and signal through FcγRIIIa (CD16) was measured as a proxy for ADCC. For spike assays, HEK293T cells were transfected with 5μg of SARS-CoV-2 original variant spike (D614G), Beta or Delta spike plasmids using PEI MAX 40,000 (Polysciences) and incubated for 2 days at 37°C. Expression of spike was confirmed by differential binding of CR3022 and P2B-2F6 and their detection by anti-IgG APC staining measured by flow cytometry. Spike transfected cells (1×10^5^ per well) were incubated with heat inactivated plasma (1:100 final dilution) or monoclonal antibodies (final concentration of 100 μg/mL) in RPMI 1640 media supplemented with 10% FBS 1% Pen/Strep (Gibco, Gaithersburg, MD) for 1 hour at 37°C. Jurkat-Lucia(tm) NFAT-CD16 cells (Invivogen) (2×10^5^ cells/well) were added and incubated for 24 hours at 37°C, 5% CO_2_. Twenty microliters of supernatant was then transferred to a white 96-well plate with 50 μl of reconstituted QUANTI-Luc secreted luciferase and read immediately on a Victor 3 luminometer with 1s integration time. The RLU of a no antibody control was subtracted as background. Palivizumab was used as a negative control, while CR3022 was used as a positive control, and P2B-2F6 to differentiate the Beta from the D614G variant. RLUs for spikes were normalised to each other and between runs using CR3022. To induce the transgene, 1X cell stimulation cocktail (Thermofisher Scientific, Oslo, Norway) and 2 μg/mL ionomycin in R10 was added to confirm sufficient expression of the Fc receptor.

### Statistical analysis

Analyses were performed in Prism (v9; GraphPad Software Inc, San Diego, CA, USA) and graphs generated using the ggplot2 package in R version 4.1.0. Non-parametric tests were used for all comparisons and all t tests were 2-sided. The Mann-Whitney test was used for unpaired comparisons between two groups, while the Kruskal-Wallis ANOVA with Dunns correction was used for multiple comparisons for unpaired groups. The Friedman test was used for multiple comparisons between paired groups. *P* values less than 0.05 were considered to be statistically significant.

## Notes

### Competing Interest Statement

The authors have declared no competing interest.

### Funding Statement

This study was funded by the South African Medical Research Council (grants 96825 and 96838). P.L.M. is supported by the South African Research Chairs Initiative of the Department of Science and Innovation and the National Research Foundation of South Africa (grant no. 98341). W.A.B. is supported by the EDCTP2 programme of the European Unions Horizon 2020 programme (TMA2016SF-1535-CaTCH-22), the Wellcome Centre for Infectious Diseases Research in Africa (CIDRI-Africa) which is supported by core funding from the Wellcome Trust (203135/Z/16/Z) and the Poliomyelitis Research Foundation (PRF 21/65). N.A.B.N acknowledges funding from the SAMRC, MRC UK, NRF and the Lily and Ernst Hausmann Trust. S.I.R. is a LOreal/UNESCO Women in Science South Africa Young Talents awardee. Related research by the authors is conducted as part of the DST-NRF Centre of Excellence in HIV Prevention, which is supported by the South African Department of Science and Technology and the National Research Foundation.

### Author Declarations

Human Research Ethics Committees of the University of the Witwatersrand (ethics reference number: M210465), University of Pretoria (ethics reference number: 247/2020) and University of Cape Town (ethics reference numbers: 190/2020 and 209/2020) gave ethical approval for this work.

### Summary of Updates

Additional neutralization data against the Omicron variant for the breakthrough infection and non-breakthrough participants at 6 months post-vaccination was added to the manuscript.

