## Supplementary Information for "Ad26.COV2.S breakthrough infections induce high titers of neutralizing antibodies against Omicron and other SARS-CoV-2 variants of concern"

### Table S1: Study participant details

| PTID[a] | Month of Vaccination | Age range at Vaccination | Sex | Month of BTI[b] | Severity of infection |
|---|---|---|---|---|---|
| BTI01 | 2021/02 | 56-60 | F | 2021/06 | Mild |
| BTI02 | 2021/02 | 36-40 | F | 2021/06 | Mild |
| BTI03 | 2021/02 | 36-40 | F | 2021/07 | Mild |
| BTI04 | 2021/02 | 56-60 | F | 2021/07 | Mild |
| BTI05 | 2021/03 | 31-35 | F | 2021/08 | Mild |
| BTI06* | 2021/02 | 31-35 | M | 2021/07 | Mild |
| V01 | 2021/03 | 56-60 | F | N/A | N/A |
| V02 | 2021/03 | 31-35 | F | N/A | N/A |
| V03 | 2021/03 | 31-35 | M | N/A | N/A |
| V04 | 2021/03 | 26-30 | F | N/A | N/A |
| V05 | 2021/03 | 31-35 | M | N/A | N/A |
| V06 | 2021/03 | 31-35 | F | N/A | N/A |
| V07 | 2021/03 | 51-55 | F | N/A | N/A |
| V08 | 2021/03 | 26-30 | F | N/A | N/A |
| V09 | 2021/03 | 31-35 | F | N/A | N/A |
| V10 | 2021/03 | 26-30 | F | N/A | N/A |
| V11 | 2021/03 | 26-30 | F | N/A | N/A |
| V12 | 2021/03 | 36-40 | F | N/A | N/A |
| V13 | 2021/03 | 26-30 | F | N/A | N/A |

[a] Participant identifier (PTID)

[b] Breakthrough infection (BTI)

* No longitudinal samples available, only 1-month post-BTI sample

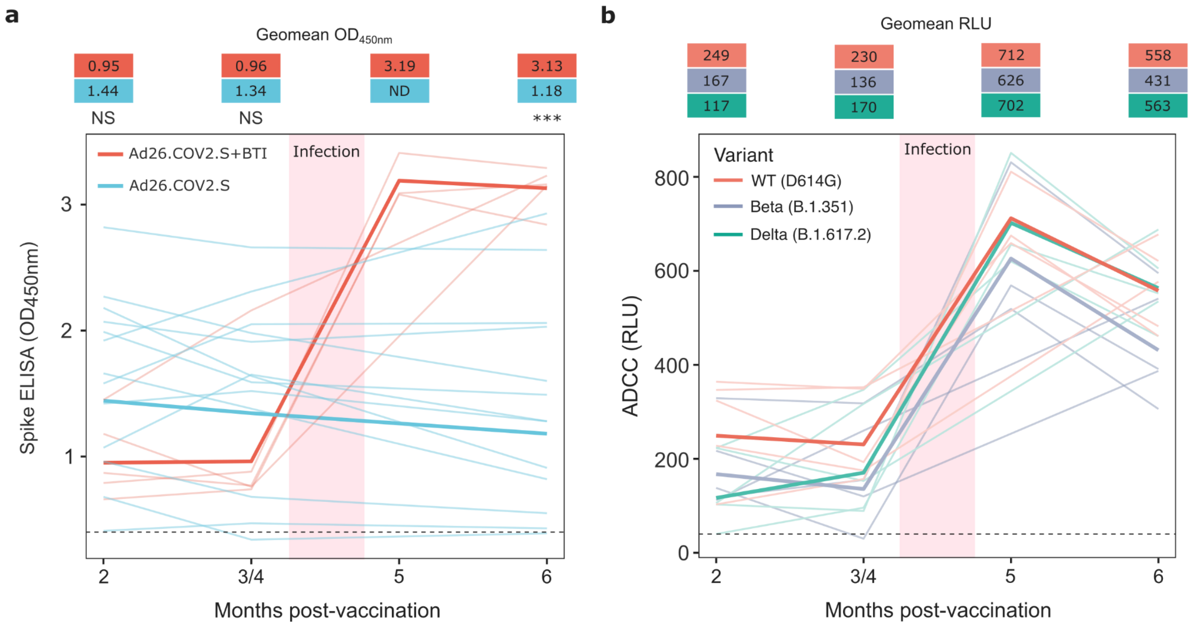


**Fig. S1: Spike specific antibody responses and antibody-dependent cellular cytotoxicity (ADCC) activity up to 6-months post-vaccination. (a)** Plasma samples from BTI and non-BTI participants were assessed for binding to the D614G spike protein ($OD_{450nm}$) by ELISA. Spike binding responses for BTI and non-BTI participants are shown in red and blue, respectively, with each line showing the longitudinal response of an individual participant over time. Lines in bold show the geomean $OD_{450nm}$ value for each group. The threshold for positivity is indicated by a dashed line. Statistical analyses were performed using the Mann-Whitney test between groups, with *** denoting $p<0.001$, NS for non-significant and ND for no data. **(b)** Cross-reactive ADCC activity for BTI participants against the D614G, Beta and Delta variants up to 6 months post-vaccination, shown as relative light units (RLUs). The threshold for positivity is indicated by a dashed line.

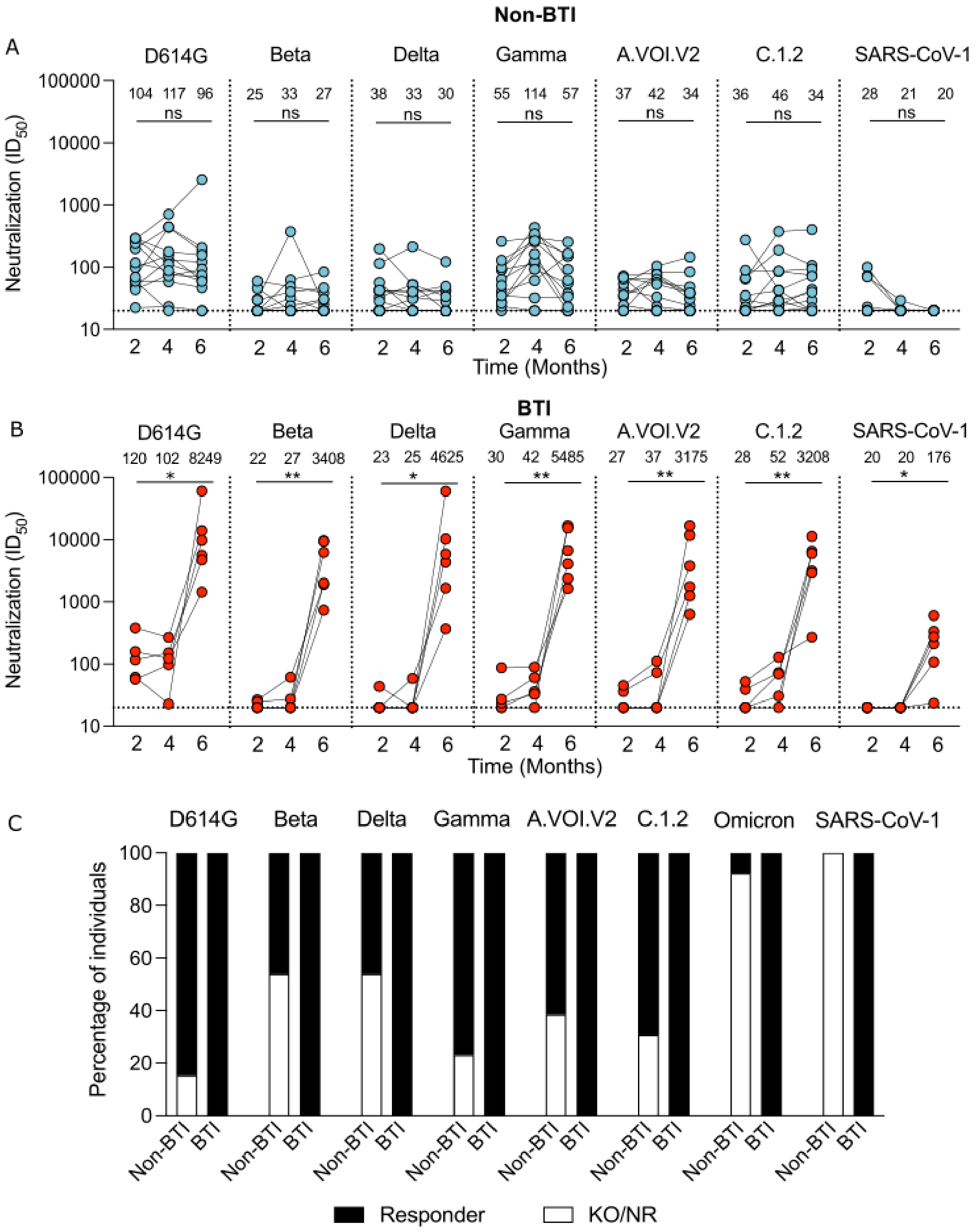


**Fig. S2: Longitudinal neutralization responses over 6 months for BTI and non-BTI participants against SARS-CoV-2 variants and SARS-CoV-1.** Neutralization $ID_{50}$ titers are shown for **(a)** Ad26.COV2.S vaccinees that did not have breakthrough infection (non-BTI) and **(b)** BTI Ad26.COV2.S vaccinees at 2-, 3- or 4- and 6-months, against D614G, Beta, Delta, Gamma, C1.2. and A.VOI.2 variants, and SARS-CoV-1. Significance is shown as per Friedman test, across all time points where ns denotes non-significant, $*p<0.05$; $**p<0.01$. **(c)** Percentage of individuals who are neutralization responders (Black; $ID_{50}$ >20), or are either non-responders or show knock-out relative to D614G (KO/NR, $ID_{50}$ <20; white) at 6-months post-vaccination.

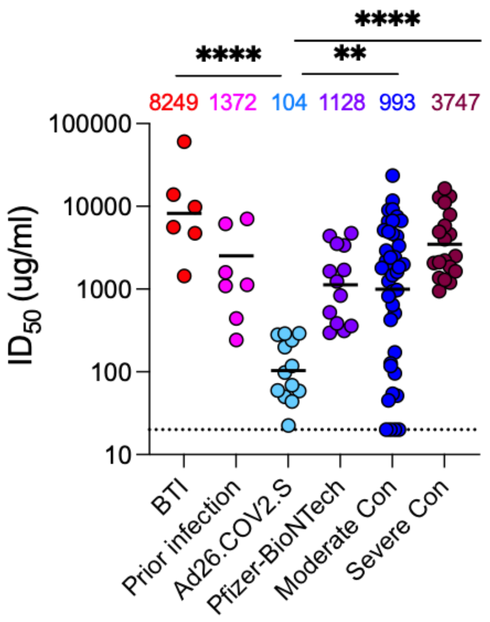


**Fig. S3: BTI neutralization responses to the D614G variant compared to vaccinees and convalescent plasma.** Neutralization $ID_{50}$ titers of two-dose Pfizer-BioNTech (2-months post-vaccination) and one-dose Ad26.COV2.S vaccinees (2-months post-vaccination) and hospitalised convalescent individuals with moderate and severe infection 10-days after a positive PCR test are shown compared to BTI individuals (1-month post-infection). Individuals who were previously infected by SARS-CoV-2 prior to Ad26.CoV2.S vaccination are shown 2-months post-vaccination. Limit of detection is indicated by a dotted line. GMT is indicated by the black bars and above the plot, while significant differences between groups, as calculated by Kruskal-Wallis ANOVA with Dunns correction, are shown above the graph with with ** denoting p<0.01 and **** denoting p<0.0001.
